# Substance use disorders exhibit unique and disorder specific genetic associations with externalizing and internalizing psychopathology

**DOI:** 10.64898/2026.01.19.26344398

**Authors:** Holly E. Poore, Maia Choi, David Zald, Denise A. Hien, COGA Collaborators, Peter B. Barr, Danielle M. Dick

## Abstract

**Background and Aims:** Substance use disorders (SUDs) are heritable and share genetic variance with externalizing and internalizing psychopathology. Although recent gene identification efforts have demonstrated the value of modeling the shared genetic architecture among SUDs and externalizing, most research has thus far failed to account for overlap with internalizing. In this study, we aim to characterize the genetic relationships of both externalizing and internalizing with SUDs.

**Design and setting:** We used genome-wide association study (GWAS) summary statistics derived from previously published studies of externalizing, internalizing, and SUD outcomes to quantify the genetic overlap between these phenotypes. We characterize this overlap using omnibus, partial, and local genetic correlations, estimates of their shared polygenic effects, genetic causality models, polygenic score (PGS) analyses, and estimates of each SUDs residual variance derived from models in Genomic SEM.

**Participants:** We used GWAS summary statistics from individuals whose genomes were most similar to those from reference panels sampled from Europe (*N*s ranged from 45,395 to 1,565,618) and Africa (*N*s ranged from 30,000 to 122,571). For polygenic scores analyses, we used data from individuals of European and African ancestry groups available in the Collaborative Study on the Genetics of Alcoholism (COGA) sample (*N*_*Maximum*_ = 7,394 for European-like genomes and 3,238 for African-like genomes)

**Measurements:** Measurements in this study include GWAS summary statistics for externalizing, internalizing, and four substance use disorders: problematic alcohol use (PAU), cannabis use disorder (CUD), opioid use disorder (OUD), and tobacco use disorder (TUD). SUD outcomes in COGA were DSM-IV symptom counts of AUD, CUD, and OUD and scores on the Fagerstrom Test for Nicotine Dependence.

**Findings:** We found strong genetic relationships of externalizing and, to a lesser extent, internalizing with all SUDs across methods. Despite their more modest associations, internalizing emerged as an important genetic correlate of SUDs. After accounting for variance shared with externalizing, partial genetic correlations between internalizing and SUDs were attenuated but, with the exception of TUD, still significant. Similarly, the PGS_INT_ accounted for a statistically significant increase in variance over and above PGS_EXT._ Two SUD specific patterns emerged such that TUD was least associated with both psychopathology spectra and OUD was most strongly related to internalizing relative to other SUDs.

**Conclusions:** From these findings we conclude that shared genetic influences may explain comorbidity observed between SUDs and internalizing disorders and suggest that genetic risk for internalizing should be incorporated into SUD identification and prevention efforts. Future gene identification efforts should study SUDs in the context of both externalizing and internalizing psychopathology.

---

Decades of twin research indicates that substance use disorders (SUDs) are moderately heritable (~40-60%) [1]. More recently, genome-wide association studies (GWAS) have begun to identify the specific genetic variants associated with the development [2, 3] of SUDs. Gene identification studies have traditionally conceptualized SUDs as isolated constructs that are separate from other forms of psychopathology; however, more recent work has found that modeling genetic covariation shared between SUDs and other psychiatric disorders can improve power and specificity of genetic associations [4].

To date, work modeling this covariation has focused on the joint risk of SUDs (e.g., broad addiction risk) [5, 6] and influences shared with other traits related to behavioral disinhibition and antagonism (e.g., externalizing spectrum) [7]. Indeed, modeling genetic influences on SUDs alongside these related phenotypes has resulted in increased power to detect genetic influences on SUDs and facilitated characterization of shared and drug specific genetic effects [4, 8, 9]. This work is an important step towards improving the precision of our targets in gene identification for SUDs, which leads to better powered and more precise biological insights [4]. Nevertheless, to date, these studies have not accounted for potential genetic overlap of SUDs with other forms of psychopathology, particularly mood and anxiety disorders, with which SUDs are often comorbid [10]. Resolving and modeling these patterns of genetic covariation with both forms of psychopathology is an important next step to identify specific neurobiological mechanisms that underpin SUDs and improve the translational utility of genetic research.

In the current study, we sought to characterize the genetic relationships between SUDs and two broad spectra of psychopathology: 1) internalizing, which encompasses traits and disorders characterized by negative emotionality, such as anxiety and depression, and 2) externalizing. We focus on these two spectra for three reasons. First, models of addiction typically highlight the importance of traits related to both spectra [11].One such model posits that genetically influenced traits of reward related risk taking (a trait related to externalizing) and negative affect (a trait related to internalizing), as well as drug-specific pathways, explain the initiation, course, and maintenance of substance use disorders [12]. Second, SUDs are often comorbid with internalizing disorders [13, 14] and their co-occurrence is associated with distinct patterns of use [15], response to treatment [16], and prognosis for both SUD and the corresponding disorders [17, 18]. In addition, the limited research characterizing their genetic overlap does suggest shared genetic influences [19] and significant polygenic prediction whereby genetic risk for anxiety explains progression from alcohol initiation to problematic use [20]. Finally, although previous studies have already demonstrated substantial genetic overlap between externalizing and SUDs [8, 21], we include this spectrum here to facilitate direct comparison of the degrees of overlap between both psychopathology spectra and SUDs, and to study the shared and unique influences of each psychopathology spectra on different SUDs.

In this study, we quantified the degree of genetic overlap of externalizing and internalizing spectra with four SUDs: problematic alcohol use (PAU), cannabis use disorder (CUD), opioid use disorder (OUD), and tobacco use disorder (TUD). Our choice to operationalize externalizing and internalizing as broad spectra reflects the greater statistical power that results from multivariate, relative to univariate, GWAS and evidence that relationships between individual spectra indicators and other traits operate through the broad dimensions [4, 8]. Conversely, we retain individual SUDs to facilitate comparison of results across different types of use disorders in light of evidence that SUDs manifest differential relationships with other forms of psychopathology [17, 22]. We use multiple methods to quantify this overlap, including genetic correlations, estimates of the polygenicity of each phenotype and overlap thereof, polygenic scores, and genetic causality analyses. Using this combination of methods, we characterize: 1) the degree of genetic overlap between internalizing and SUDs, 2) the relative contributions of externalizing and internalizing to SUDs, and 3) differential patterns of associations across the individual SUDs.

## Methods

This research was reviewed and approved the Institutional Review Board at Rutgers University (Pro2022000138).

### Samples

GWAS summary statistics were derived from samples of individuals whose genomes were most similar to those from reference panels sampled from Europe and Africa (hereafter referred to as “EUR and “AFR”; see Supplementary Table 1). Analyses were limited to these populations due to data availability.

GWAS summary statistics corresponded to phenotypes reflecting broad externalizing and internalizing psychopathology spectra and four SUDs for which well-powered GWAS were available: PAU [3], CUD [23], OUD [24], and TUD [2]. Briefly, the GWAS for CUD, OUD, and TUD reflect case-control GWAS in which cases were individuals who met ICD or DSM criteria for the disorder. The PAU GWAS is a meta-analysis of case-control GWAS of AUD and the AUDIT Problems scale [25]. Additional information about phenotyping is available in the respective GWAS publications.

EUR externalizing summary statistics were drawn from a reduced model of a recent GWAS of externalizing [8] from which the PAU indicator was removed. Although PAU was a strong indicator of externalizing in the original model [19], we removed it to prevent artificially inflating the associations between externalizing and alcohol problems in the current analyses. Unfortunately, the only non-SUD externalizing indicator available in AFR samples was a GWAS of smoking initiation [26], defined as ever having been a regular smoker, which we investigated this as a proxy for externalizing. The cross-ancestry correlation [27] between EUR externalizing and AFR smoking initiation was statistically indistinguishable from 1 (rg = 1.10, SE =.23), so we proceeded with this plan.

Finally, for the purposes of this study, we fit an internalizing model comprised of anxiety [28], major depressive disorder [29, 30], posttraumatic stress disorder [31], and neuroticism [32] for EUR and AFR samples separately in Genomic SEM [33]. We chose these phenotypes because they have shown to be strong indicators of an internalizing factor in phenotypic models [34] and well-powered GWAS of these phenotypes were available in both EUR and AFR samples. We estimated single nucleotide polymorphism (SNP) effects on each factor and performed multi-marker analysis of genomic annotation (MAGMA; version 1.08) [35] to identify genes associated with internalizing as well as gene-property analysis of enrichment in GTEx v8 [36] tissue types using Functional Mapping and Annotation of Genome-Wide Association Studies (FUMA) version 1.6.1 [37]. See Supporting Information for additional detail.

## Statistical Analyses

To account for differences in linkage disequilibrium (LD) structure and minor allele frequency differences, all analyses were conducted separately in EUR and AFR samples, unless otherwise noted. We used six methods to characterize the relationships between individual SUDs and psychopathology spectra (i.e., externalizing and internalizing). Some analyses were used to quantify the relationship between individual SUDs and a single spectrum at a time (e.g., OUD with externalizing), whereas others allowed us to examine the relationship between individual SUDs and both spectra (e.g., a joint analysis of OUD with externalizing and internalizing). The latter analyses provide information about the relative association of an SUD with one spectrum while accounting for its relationship with the other (e.g., the correlation between OUD and internalizing after accounting for externalizing) and the degree of unique variance each SUD retains after accounting for both spectra. These methods are detailed below.

### Genetic correlations

We first estimated zero-order and partial genetic correlations (partialling out genetic variance of the other psychopathology spectra) using the *partialLDSC* package in R [38]. Partial genetic correlations reflect the correlation between the psychopathology spectrum and the SUD after accounting for the shared variance between the two spectra (e.g., correlation between externalizing and OUD after accounting for internalizing). Differences in the zero-order and partial correlations show the degree to which shared variance between externalizing and internalizing explains the relationships between individual spectra and the SUD. We next used Local Analysis of (co)Variant Association (LAVA) [39] to estimate local genetic correlations between the individual spectra and each SUD at 2,495 loci across the genome. This analysis facilitates investigation of potential differential direction or strength of effects that may occur at specific segments of the genome. We applied a within-ancestry Bonferroni *p*-value correction to determine significance of local genetic correlations.

### Polygenic overlap

Bivariate MiXeR [40, 41] was used to estimate the degree of polygenic overlap between pair-wise combinations of each psychopathology spectra and SUD. MiXeR applies gaussian mixture models to estimate the proportion of genetic variants that influence both traits, one trait but not the other, and have no influence on either trait. Output includes a Dice coefficient, which is an index of similarity calculated as the proportion of polygenic variants versus total number of influential variants for both traits, as well as r_gs,_ which is the correlation of effect sizes within the shared polygenic component. Finally, it provides fit statistics to distinguish between different models of the degree of overlap (e.g., minimum, polygenic, maximum) between the two traits. Due to concerns about power requirements, MiXeR was only applied to EUR samples.

### Genetic causality

Latent causal variable (LCV) modeling [42] was used to test the hypothesis of genetic causality between each psychopathology spectrum and SUD. In an LCV model, the genetic correlation between two traits is mediated by a latent variable that has a causal effect on each trait. The extent to which trait one (e.g., externalizing) is correlated with the latent variable can be used to estimate the degree to which trait one is genetically causal for trait two (e.g., PAU). Output from LCV includes a genetic causality proportion (GPC), which is an estimate of this causal relationship and ranges from −1 to 1, with 0 indicating no genetic causality and the direction of effect indicating the direction of causal effects. In accordance with the authors’ recommendations, we only interpreted results from models in which the Z score of the heritability estimate for both traits was > 7.

### Polygenic scores

Polygenic scores (PGS) for EXT and INT were computed using PRS-CSx, which uses a shared continuous shrinkage parameter to incorporate information from GWAS of different ancestry populations to improve effect size estimation, and estimated their associations with SUD outcomes in the Collaborative Study on the Genetic of Alcoholism (COGA) study. Briefly, COGA [43] is a multi-site family-based sample of individuals in treatment for alcohol use disorder and their families as well as community-based comparison families. SUD outcomes were symptom counts of AUD, CUD, and OUD [44] and scores from the Fagerstrom Test of Nicotine Dependence (FTND) [45] obtained through semi-structured interviews (i.e., the SSAGA [44]). Sample sizes available for each variable ranged from 1,700 (OUD symptoms) to 7,394 (AUD symptoms) in EUR participants and 533 (OUD symptoms) to 3,238 (AUD symptoms) in AFR participants. We used least squares regression and adjusted the standard errors in COGA to account for family structure. Regression models were estimated using the *estimatr* package in R [46]. We evaluated the incremental R^2^ (Δ*R*^2^) attained by adding the polygenic scores to a regression with baseline covariates (e.g., age, sex, and ancestry PCs). We estimated 95% CIs for Δ*R*^2^ using bootstrapping (1,000 iterations) in the *boot* R package [47]. We ran models in which each SUD outcome was regressed on both PGS and covariates and compared the Δ*R*^2^ in these models to the Δ*R*^2^ which included only one PGS at a time.

### Genomic SEM models

We estimated the proportion of unique genetic variance each SUD retained after accounting for what it shared with externalizing and internalizing in Genomic SEM [33]. We did this by fitting a series of trivariate Cholesky decomposition models (see Figure 1) that included internalizing, externalizing, and one SUD (e.g., PAU) at a time. The first factor included internalizing, externalizing, and one SUD, to account for what all three variables share. The second factor included only externalizing and the SUD, to account for what’s shared between them that isn’t also shared with internalizing. Finally, only the SUD loaded on the last factor, which represents its residual variance leftover after accounting for internalizing and externalizing.

**Figure 1.**
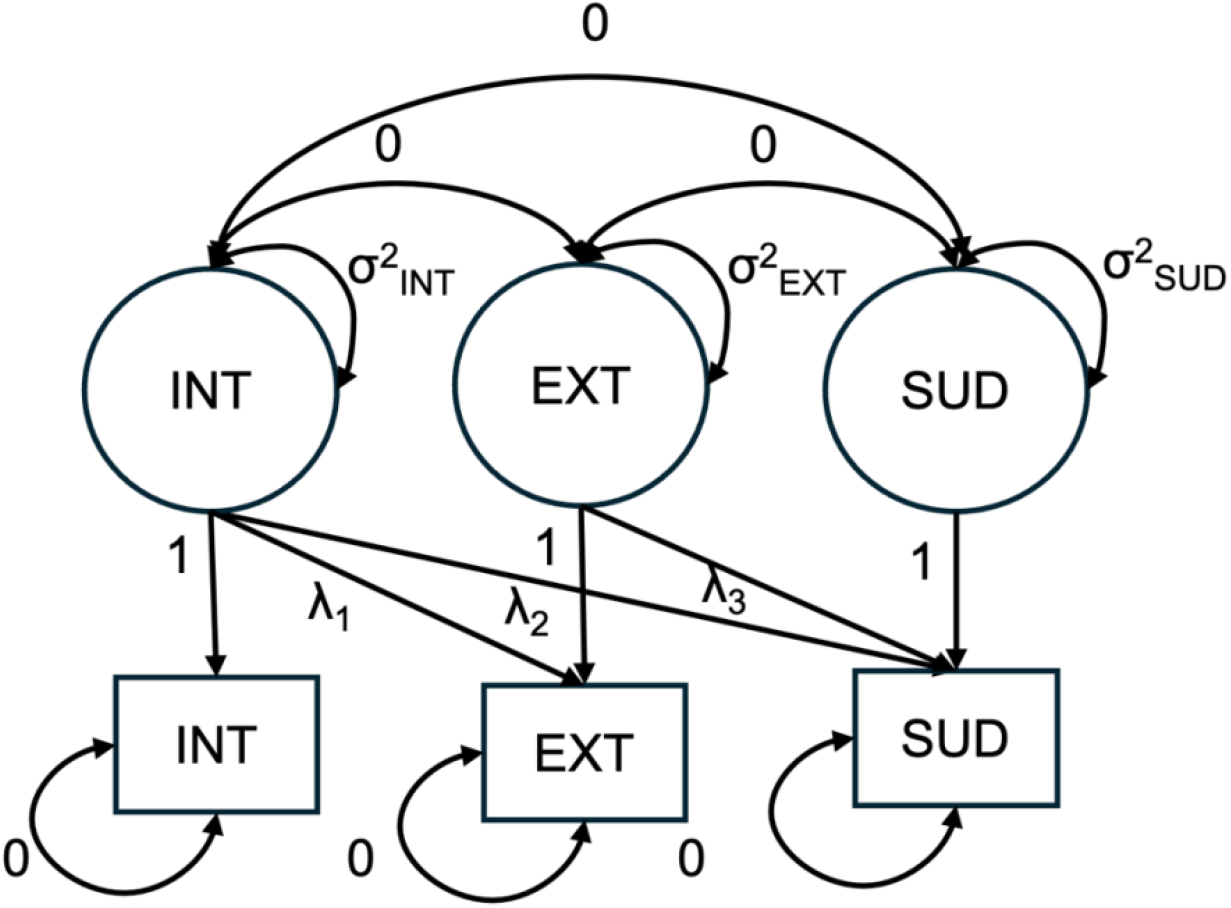
Example trivariate Cholesky models. These models were used to estimate the residual variance of each SUD (PAU, CUD, OUD, and TUD) after accounting for what is shares with internalizing and externalizing. INT = internalizing, EXT = externalizing, SUD = the SUD included in the model. Analyses were performed separately with EUR and AFR samples.

## Results

### Genetic associations between externalizing and SUDs

As expected, all SUDs were strongly genetically correlated with externalizing and this association appeared to be independent of genetic influences on internalizing, as all partial correlations remained strong and significant (Figure 2a, Supplementary Table 6). Similarly, all local genetic correlations between externalizing and SUDs in EUR samples were moderate to strong (r_g_s ranged from.30 to 1.0 and 88% of correlations >.50) and evenly distributed across the genome (Figure 2b, Supplementary Table 7). No local correlations between psychopathology spectra and any SUD were statistically significant after applying a p-value correction in the AFR samples (Supplementary Table 8). In bivariate MiXeR models, we observed high degrees of overlap between externalizing and each SUD (Figure 1c; Supplementary Table 9), with model fit statistics for CUD and OUD supporting a maximal overlap model (i.e., a model in which the influential variants for each SUD are subsumed in those for externalizing). Finally, we found evidence of a partial genetic causality relationship between externalizing and TUD in both the EUR and AFR samples (GCP_EXT-TUD_ = −0.57 (.09), p <.001 in EUR and GCP_EXT-TUD_ = −0.81 (.16), p <.001 in AFR; Supplementary Table 10). All GPC coefficients were negative, indicating that the genetic influences on TUD were causal for the psychopathology spectra (rather than that genetic influences for the spectra were causal for TUD).

**Figure 2.**
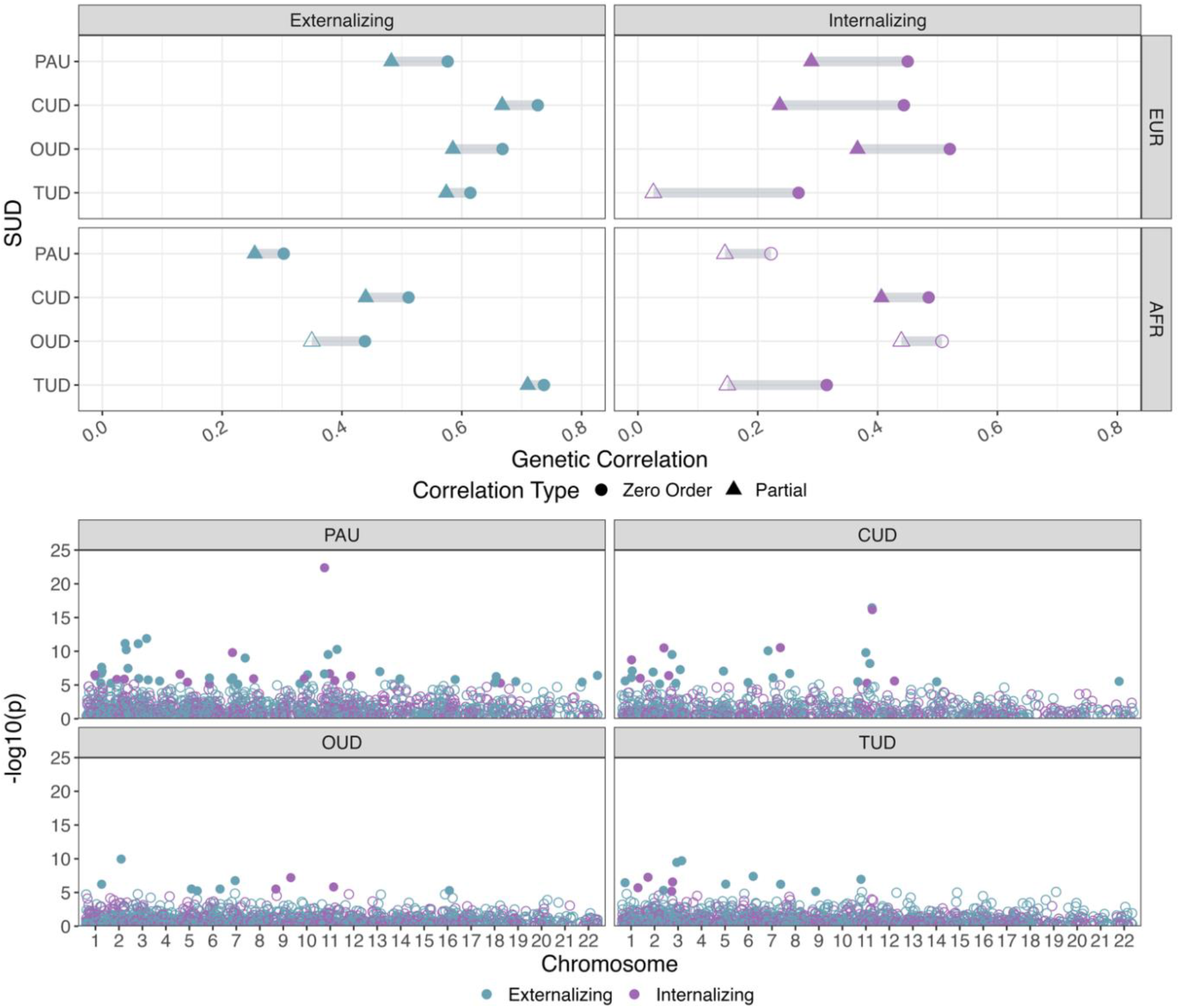
Genetic correlations between psychopathology spectra and SUDs. (**top**) zero-order (circle) and partial (triangle) correlations of SUDs with psychopathology spectra for EUR- and AFR-like samples. Filled in shapes indicate correlations significant at *p* <.05 whereas open shapes indicate correlations that were not significant. **(bottom)** local genetic correlations of each SUD with externalizing and internalizing across the genome in EUR-like samples. AFR results are not shown because no local correlations between psychopathology spectra and any SUD were statistically significant after applying a p-value correction. Filled in shapes indicate correlations with a Bonferroni corrected *p*-value <.05 whereas open shapes indicate a nonsignificant *p*-value.

**Figure 3.**
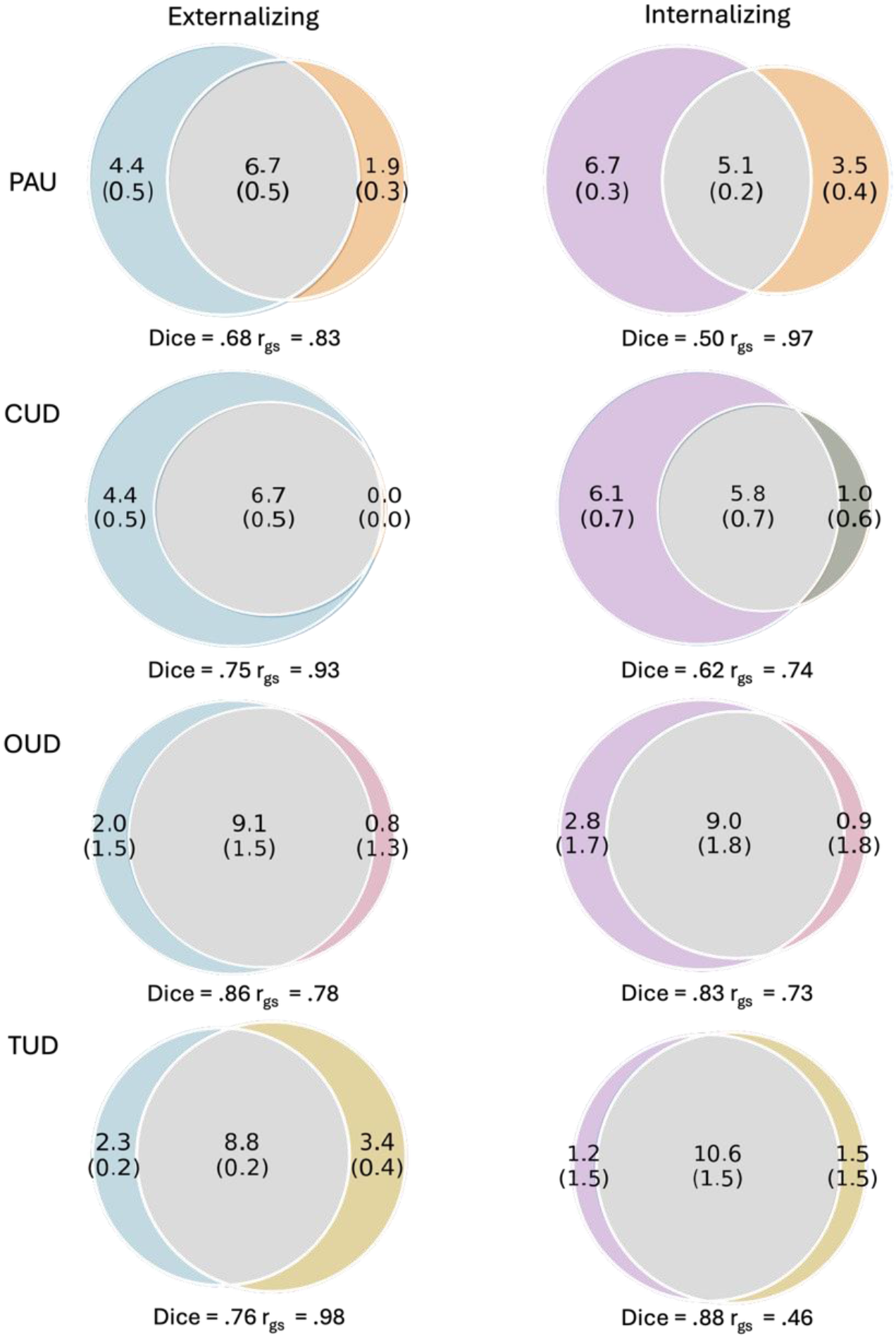
**MiXeR results** as shown by venn diagrams representing polygenic overlap of externalizing and internalizing with SUDs. The number of influential variants is shown (in thousands) in blue for externalizing and purple for internalizing. The number of estimated shared influential variants for each pair of traits is shown in gray. Numbers in parentheses are the standard errors (in thousands) of these estimates. Dice = an index of similarity ranging from 0-100, calculated as the proportion of polygenic variants versus total number of influential variants for both traits. r_gs_ = the correlation of effect sizes within the shared polygenic component.

**Figure 4.**
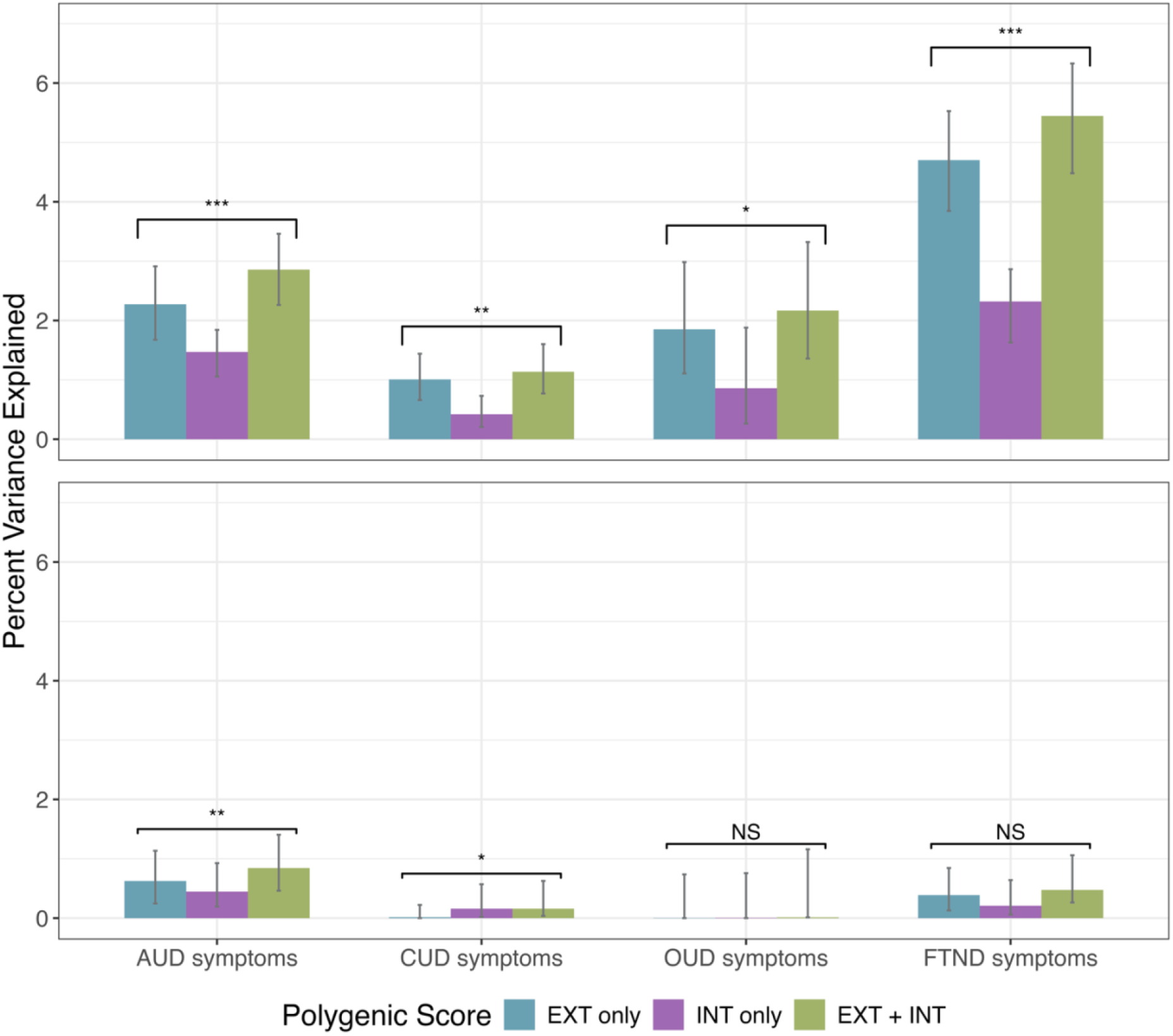
Percent variance explained by polygenic scores in SUD outcomes. Percent of variance accounted for by externalizing (purple), internalizing (blue), and externalizing and internalizing (green) polygenic scores (PGS) in SUD outcomes in COGA. Asterix indicate the significant incremental variance explained when the internalizing PGS is added to a model with the externalizing PGS. *** indicates *p* <.001, ** indicates *p* <.01, * indicates *p* <.05, NS indicates *p* >.05.

### Genetic associations between internalizing and SUDs

SUDs were weakly to moderately correlated with internalizing. Although these associations were more attenuated when accounting for externalizing, many remained statistically significant (Figure 1a, Supplementary Table 6). The exception to this was the partial correlation of internalizing with TUD, which was no longer significant. These results are largely consistent across ancestry groups, although lower power in the AFR samples contributed to higher standard errors, limiting interpretation of these results (Supplementary Table 6). We found overall fewer local genetic correlations between internalizing and SUDs, but most correlations that survived p-value correction were moderate to strong (ranging from −.81 to.95, with 76% of r_g_s > |.50|, Figure 1b, Supplementary Tables 7-8). The one negative correlation was with PAU on a locus on Chromosome 12. In addition, correlations of internalizing with OUD and TUD were somewhat more localized relative to other SUDs, with their significant correlations occurring in loci between chromosomes 9 to 11 and 1 to 3, respectively. Interestingly, the region on chromosome 11 contains *NCAM1*, which has been separately associated with several substance use disorders [48–50], broad externalizing [8], and internalizing traits (e.g., neuroticism, PTSD) [31, 51] the regions on chromosome 3 contain several genes previously implicated in smoking and anxiety, or mood related phenotypes (e.g., *SLC38A3, ROBO2)* [28, 48, 51, 52]. In MiXeR analyses, we observed particularly strong overlap between internalizing and OUD, with model fit criteria again supporting a maximal overlap model (Figure 1c, Supplementary Table 9). Finally, LCV analyses supported a genetic causality relationship between internalizing and TUD (GCP_INT-TUD_ = −0.17 (.05), p <.001), indicating that TUD is partially genetically causal for internalizing (Supplementary Table 10).

### Genetic associations between both psychopathology spectra and SUDs

PGS_EXT_ accounted for 1.01% (CUD symptoms) to 4.70% (FTND symptoms) of the variance in SUD symptom dimensions in EUR-like individuals in COGA and between 0% (OUD symptoms) and 0.62% (AUD symptoms) of the AFR-like individuals (Figure 1e, Supplementary Table 11). PGS_INT_ accounted for 0.86% (OUD symptoms) to 2.32% of the variance (FTND symptoms) in EUR-like participants and 0.01% (OUD symptoms) to 0.45% (AUD symptoms) in AFR-like participants. Despite this more modest association, the addition of PGS_INT_ in a model with PGS_EXT_ resulted in a statistically significant increase in variance accounted for in all SUD symptom dimensions in EUR-like participants (incremental R^2^ ranged from 0.13% [CUD symptoms] to 0.74% [FTND symptoms]) and in AUD (incremental R^2^ = 0.12%) and CUD symptoms (incremental R^2^ = 0.14%) in AFR-like participants (Supplementary Table 10).

Finally, residual variance estimates for each SUD in the trivariate Cholesky models (i.e., genetic variance unique from externalizing and internalizing) were highest for TUD and CUD (.62 [.05] and 0.61 [.05], respectively) followed by PAU (0.48 [.10) and OUD (0.45 [.05]) in EUR samples. In AFR samples, PAU retained the most residual variance (0.89 [.16), followed by OUD (0.65 [.67]), CUD (0.62 [.16]), and TUD (0.45 [.16]).

## Discussion

In the current study, we used multiple, complimentary genetic association methods to quantify the degree of overlap of externalizing and internalizing with individual SUDs in EUR and AFR ancestry samples. As expected, we found strong genetic relationships between externalizing and each of the SUDs studied here across methods. This is consistent with a longstanding literature supporting the inclusion of SUDs in the externalizing spectrum and evidence of shared phenotypic and genetic overlap [7, 53], so much so that the joint analysis of these phenotypes has been shown to increase power to detect genetic effects for SUDs [4]. We also found evidence for appreciable, though more modest, genetic associations between internalizing and SUDs. These associations remained significant even after accounting for shared variance between internalizing and externalizing, supporting a unique role of internalizing in characterizing genetic influences on SUDs. Our findings suggest that phenotypic comorbidity observed between internalizing disorders and SUDs may be driven, in part, by pleiotropic genetic influences that give rise to both phenotypes. This underscores the importance of considering negative affect in early identification, prevention, and treatment efforts and supports including measurement of genetic risk for internalizing, in addition to externalizing [54] in these efforts.

The differences in findings across externalizing and internalizing broadly is worth noting. Relative to externalizing, internalizing had somewhat more specific associations with respect to its pattern of genetic correlations across SUDs and the regions of the genome that give rise to these correlations. For example, whereas externalizing was uniformly associated with all SUDs, internalizing was most strongly associated OUD (discussed in more detail below) and least strongly with TUD to the point that its partial genetic correlation was nonsignificant after accounting for shared variance with internalizing. Similarly, local genetic correlations suggested that genetic correlations between externalizing and SUDs are driven by shared genetic effects distributed across the genome whereas those between internalizing and SUDs are more localized and in regions that contain genes previously associated with both SUDs and internalizing traits [31, 48, 51, 52].

Our findings also revealed two interesting patterns for specific SUDs that are worth highlighting. First, TUD appeared to be the least associated with either psychopathology spectrum. Although it had an appreciable genetic correlation with externalizing, it also had the fewest number of local genetic correlations, a nonsignificant partial genetic correlation with internalizing, and the most unique variance after accounting for what is shared with externalizing and internalizing. This is consistent with results from twin studies, which found that nicotine dependence shared a smaller proportion of its genetic variance with psychiatric disorders compared to other SUDs [55, 56]. It is also possible that TUD’s genetic correlations with other traits are influenced by its measurement. For example, one study found that the genetic correlations of nicotine dependence with other forms of psychopathology varied significantly depending on whether nicotine dependence was defined by DSM, ICD, or FTND criteria [57].Nevertheless, TUD is the only SUD for which we found evidence of a causal genetic relationship, perhaps indicating bivariate association methods are, at least partially, picking up on evidence of causal, rather than pleiotropic, genetic variance [58]. Second, OUD appeared to be more strongly associated with internalizing than other SUDs, as evidenced by their high genetic correlation and near total polygenic overlap. This is consistent with evidence that internalizing is particularly important in explaining more severe forms of problematic substance use [19, 59]. It is also possible that this association reflects the shared influence of negative affect and medical conditions that would increase the likelihood of opioid exposure and subsequent development of problematic use. Indeed, a recent study found evidence that shared genetic components between SUDs, broadly defined, and mood/anxiety disorders, were also shared with other health conditions, such as chronic pain [6]. Similarly, genetic risk for somatoform traits, including chronic pain, has been associated with use of opiates and OUD as well as mood and anxiety disorders in large biobanks [60].

These results should be interpreted in the context of a few limitations. First, although we were able to include both EUR and AFR samples, the lower power of the AFR summary statistics led to decreased precision of the analyses and may have led to spurious or nonsignificant findings. This impedes our interpretation of the results in AFR samples and any differences in the pattern of associations observed across the two ancestry groups. In addition, we used smoking initiation as a proxy for externalizing in AFR samples and, although the cross ancestry genetic correlation between smoking initiation and externalizing in EUR was one, this likely influenced patterns of results with specific SUDs, especially TUD. Second, comparisons across SUDs should also be interpreted with caution as they are ascertained from different samples, use different criteria for determining diagnoses, and have unequal power. Finally, we are limited to disorder-level SUD GWAS, which may mask more nuanced relationships between individual symptoms of SUD and associations with other forms of psychopathology [10, 61].

Overall, our findings support two main conclusions. First, we provide evidence that SUDs should be studied in the context of both externalizing *and* internalizing. Associations with internalizing tended to be more modest and we would not expect the substantial improvements in power that have been found when jointly modeling externalizing and SUDs [4]. Nevertheless, we believe that explicitly modeling this overlap in gene identification studies will facilitate 1) detection of variants that influence SUDs through internalizing pathways and 2) improvement in the specificity of associations found for SUDs. Second, when studying shared genetic influences between SUDs and other forms of psychopathology, it is important to model residual, substance-specific effects, as SUDs appear to manifest differential associations with the psychopathology spectra.

## Data Availability

The summary statistics used for the current study are from previously published studies and no new data were collected. These studies are described in the text. Some of these studies include data which have restricted access to protect the privacy of the study participants. The full sets of internalizing summary statistics generated by this study can be made available to qualified investigators who enter into an agreement with 23andMe that protects participant confidentiality and who have access to MVP data. Once these requests have been satisfied, and the manuscript has been published, investigators may request to use our summary statistics from the first author.

## Acknowledgements

This work was supported by the National Institute on Alcohol Abuse and Alcoholism (R01AA015416 to DMD, K02AA018755 to DMD, U10AA008401 to DMD, P50AA022537 to DMD, and T32AA032229 to MC) and the National Institute on Drug Abuse (K01DA059657 to HEP, R01DA050721 to DMD, T32DA55569 to MC).

The authors thank **The Externalizing Consortium**. Principal Investigators for EXT1.0: Danielle M. Dick, Philipp Koellinger, K. Paige Harden, Abraham A. Palmer. Lead Analysts: Richard Karlsson Linnér, Travis T. Mallard, Peter B. Barr, Sandra Sanchez-Roige. Significant Contributors: Irwin D. Waldman. The Externalizing Consortium has been supported by the National Institute on Alcohol Abuse and Alcoholism (R01AA015416 – administrative supplement to DMD), and the National Institute on Drug Abuse (R01DA050721 to DMD). Additional funding for investigator effort has been provided by K02AA018755, U10AA008401, P50AA022537 to DMD, as well as a European Research Council Consolidator Grant (647648 edge to Koellinger). The content is solely the responsibility of the authors and does not necessarily represent the official views of the above funding bodies. The Externalizing Consortium would like to thank the following groups for making the research possible: **23andme, Add Health, Vanderbilt University Medical Center’s biovu, Collaborative Study on the Genetics of Alcoholism (COGA), the Psychiatric Genomics Consortium’s (PGC) Substance Use Disorders working group, UK10K Consortium, UK Biobank**, and **Philadelphia Neurodevelopmental Cohort**. All code necessary to replicate this study is available upon request.

The authors thank **Million Veteran Program (MVP)** staff, researchers, and volunteers, who have contributed to MVP, and especially participants who previously served their country in the military and now generously agreed to enroll in the study.

Finally, we thank **The Collaborative Study on the Genetics of Alcoholism (COGA)**, Principal Investigators B. Porjesz, V. Hesselbrock, A. Agrawal; Scientific Director, A. Agrawal; Translational Director, D. Dick, includes ten different centers: University of Connecticut (V. Hesselbrock); Indiana University (H.J. Edenberg, T. Foroud, Y. Liu, M.H. Plawecki); University of Iowa Carver College of Medicine (S. Kuperman, A. Anderson); SUNY Downstate Health Sciences University (B. Porjesz, J. Meyers); Washington University in St. Louis (L. Bierut, A. Agrawal, S. Hartz); University of California at San Diego (M. Schuckit); Rutgers University (D. Dick, R. Hart, J. Salvatore, J. Tischfield); The Children’s Hospital of Philadelphia, University of Pennsylvania (L. Almasy); Icahn School of Medicine at Mount Sinai (A. Goate, P. Slesinger); and Howard University (D. Scott). Other COGA collaborators include: C. Holzhauer, M. Hesselbrock (University of Connecticut); D. Lai, J. Nurnberger Jr., L. Wetherill, X., Xuei, S. O’Connor, (Indiana University); J. Kramer (University of Iowa), G. Chan (University of Iowa; University of Connecticut); C. Kamarajan, A. Pandey, D.B. Chorlian, P. Barr, S. Kinreich, G. Pandey, Z. Neale, S., C. Chatzinakos, J. Zhang, Saenz deViteri, R. Christian, A. Bingly (SUNY Downstate); G. Pathak (Icahn School of Medicine at Mount Sinai); A. Anokhin, K. Bucholz, F. Dong, A. Hatoum, E. Johnson, V. McCutcheon, J. Rice, S. Saccone (Washington University); F. Aliev, Z. Pang, S. Kuo, S. Brislin, J. Moore (Rutgers University); A. Merikangas (The Children’s Hospital of Philadelphia and University of Pennsylvania); M. Gitik, NIAAA Staff Collaborator. We continue to be inspired by our memories of Henri Begleiter and Theodore Reich, founding PI and Co-PI of COGA, and also owe a debt of gratitude to other past organizers of COGA, including Ting-Kai Li, P. Michael Conneally, Raymond Crowe, and Wendy Reich, for their critical contributions. This national collaborative study is supported by NIH Grant U10AA008401 from the National Institute on Alcohol Abuse and Alcoholism (NIAAA) and the National Institute on Drug Abuse (NIDA).

