## Supporting Information for "Substance use disorders exhibit unique and disorder specific genetic associations with externalizing and internalizing psychopathology"

### Multivariate GWAS of internalizing


Methods & results

Supplementary Figure 1

Supplementary Figure 2

**Multivariate GWAS of internalizing**

We performed a multivariate genome-wide association study (GWAS) of internalizing from samples of individuals whose genomes were most similar to those from reference panels sampled from Europe and Africa (hereafter referred to as “EUR” and “AFR”). We selected four indicators from recent, well-powered GWAS of disorders and traits commonly included in internalizing models of psychopathology [1]. These phenotypes include: major depressive disorder (MDD) [2, 3], posttraumatic stress disorder (PTSD) [4], neuroticism [5], and anxiety [6]. With the exception of MDD, summary statistics from EUR and AFR ancestry samples were drawn from the same study. Additional details about the studies from which these summary statistics were derived can be found in Supplementary Table 1.

The summary statistics for the EUR samples reported in Friligkou et al were drawn from a GenomicSEM model which estimated single nucleotide polymorphism (SNP) effects on a latent factor indicated by six GWAS of anxiety from distinct cohorts. They chose this approach to account for differential weights of the individual GWAS. However, when merging SNPs across indicators, Genomic SEM applies list-wise deletion such that only the SNPs that are present in all indicators are included in the final model. This can result in a lower number of SNPs available for analysis compared with traditional meta-analysis, especially when further merging with other summary statistics, as we did for our internalizing model. We chose instead to use METAL [7] to perform a meta-analysis, which retains all SNPs present in at least one cohort, of the six anxiety cohorts included in the Friligkou et al publication, which results in a greater number of SNPs available for analysis. These meta-analyzed summary statistics were genetically correlated at .90 (.05) with the summary statistics from Genomic SEM. After merging summary statistics across all internalizing indicators, 6,299,993 SNPs were available in EUR samples.

The anxiety GWAS from AFR samples did not have a heritability estimate statistically different from 0, so we omitted this indicator from the model. After merging summary statistics across these three internalizing indicators, 8,897,495 SNPs were available in AFR samples.

We used 1000 Genomes Reference Panels for all analyses. Clumping was performed in Functional Mapping and Annotation of Genome-Wide Association Studies (FUMA) version 1.6.1[8] using an $r^2$ threshold of ≥ 0.6 to define independent significant SNPs, a second threshold of $r^2 \geq 0.1$ to define lead SNPs, and a maximum distance between LD blocks of 250kb to merge into a locus. To further characterize these genetic effects, we used multi-marker analysis of genomic annotation (MAGMA; version 1.08) [9], in which genome-wide SNPs were mapped to 18,235 protein-coding genes from Ensembl v102, and SNPs within each gene were jointly tested for association with internalizing. We evaluated Bonferroni corrected significance adjusted for the number of genes (one sided $p < 2.74\text{x}10^{-6}$). We also used MAGMA tissue expression analysis to test the relationship between expressed genes in different tissues and EUR internalizing (Supplementary Figure 2). Tissue expression analysis used weights from GTEx v8 [10]. MAGMA identified 807 genes (Supplementary Table 4) and tissue expression showed the strongest enrichment in the cerebellum and frontal cortex (Supplementary Figure 2, Supplementary Table 5).

Model fit and factor loadings for the internalizing models in EUR and AFR ancestry samples are presented in Supplementary Table 2. Briefly, model fit was good in the EUR model and factor loadings were strong and significant (standardized loadings ranged from .76 [neuroticism] to .94 [PTSD]). The AFR internalizing model only had three indicators and was just identified, model fit statistics were not available. Nevertheless, the loadings were strong and significant (standardized loadings ranged from .80 [PTSD] to .91 [neuroticism]).

Multivariate GWAS of EUR internalizing identified 469 genomic risk loci and MAGMA gene-based analyses identified 807 genes (Supplementary Figures 1-2, Supplementary Tables 3-5). Top loci include rs7111031 mapped to *DRD2* gene, which encodes the D2 subtype of the dopamine receptor and was previously associated with wellbeing and depression phenotypes [11, 12], and rs11763750, mapped to *MAD1L1* and previously associated with suicidal ideation and depression phenotypes [13, 14]. No significant SNPs were identified in the AFR GWAS.

To assess the homogeneity of SNP effects on the internalizing factor, we calculated $Q_{SNP}$ heterogeneity statistics, which can be used to identify SNPs that have an effect on one or more indicator phenotypes that is better explained by pathways independent of the factor. A large number of $Q_{SNPs}$ indicate heterogeneous factors that are not well represented by their indicators and do not cohere at the level of the genome. We identified 34 $Q_{SNP}$ loci for EUR internalizing, which is low relative to the total number of genomic risk loci identified for the factor. In addition, only one $Q_{SNP}$ loci overlapped with any of the 469 internalizing loci, indicating that identified genetic variants represent true effects on the broad factor. The one overlapping SNP was rs13262595, which is an intronic variant located in *TSNARE1* that has previously been associated with a wide variety of psychiatric disorders and traits (e.g., schizophrenia, anxiety, ADHD) in the GWAS catalog [15].

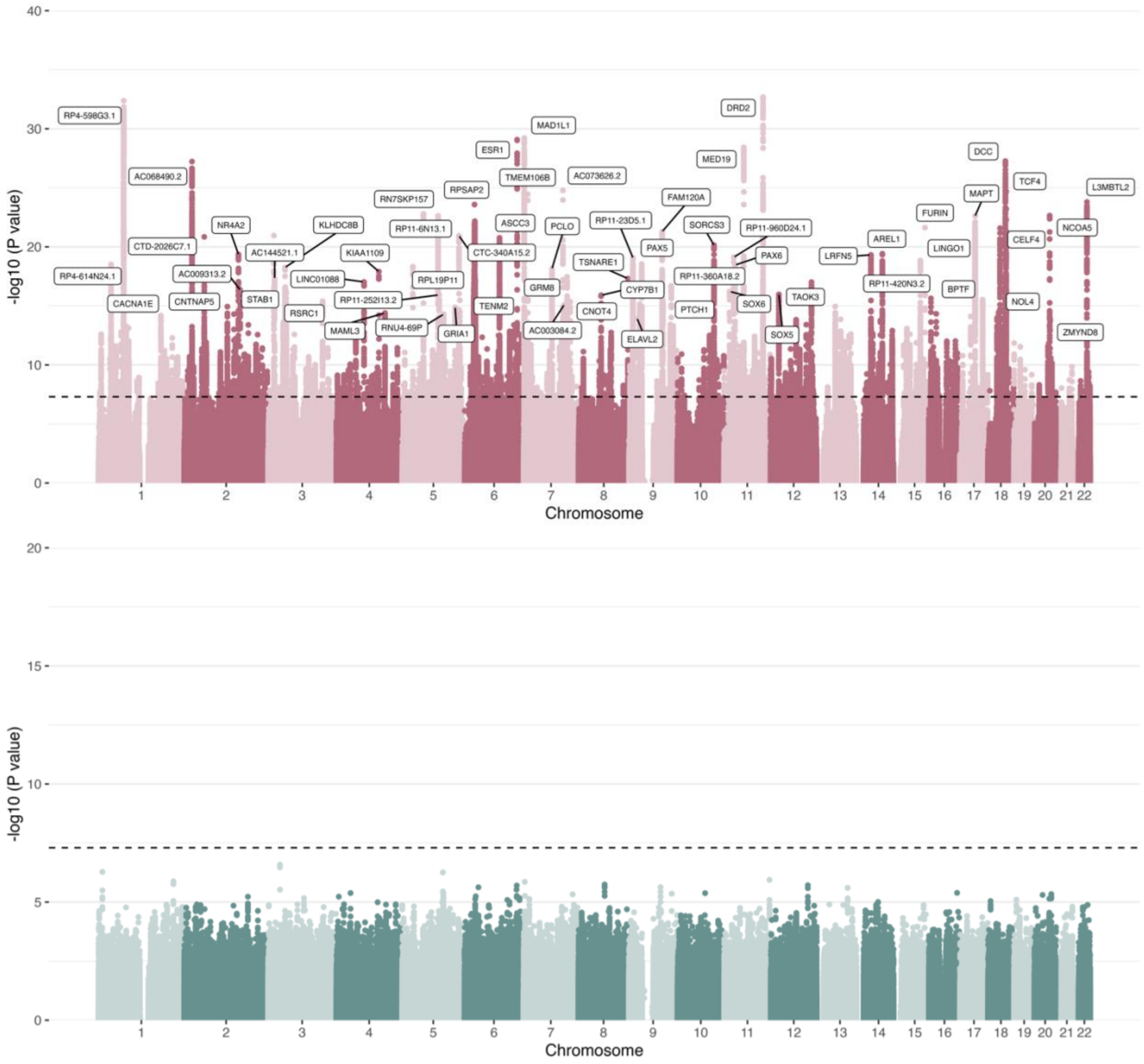


**Supplementary Figure 1.** Manhattan plots of internalizing in EUR (top) and AFR (bottom) samples. Top loci are mapped to the nearest gene using ANNOVAR [16] annotation.

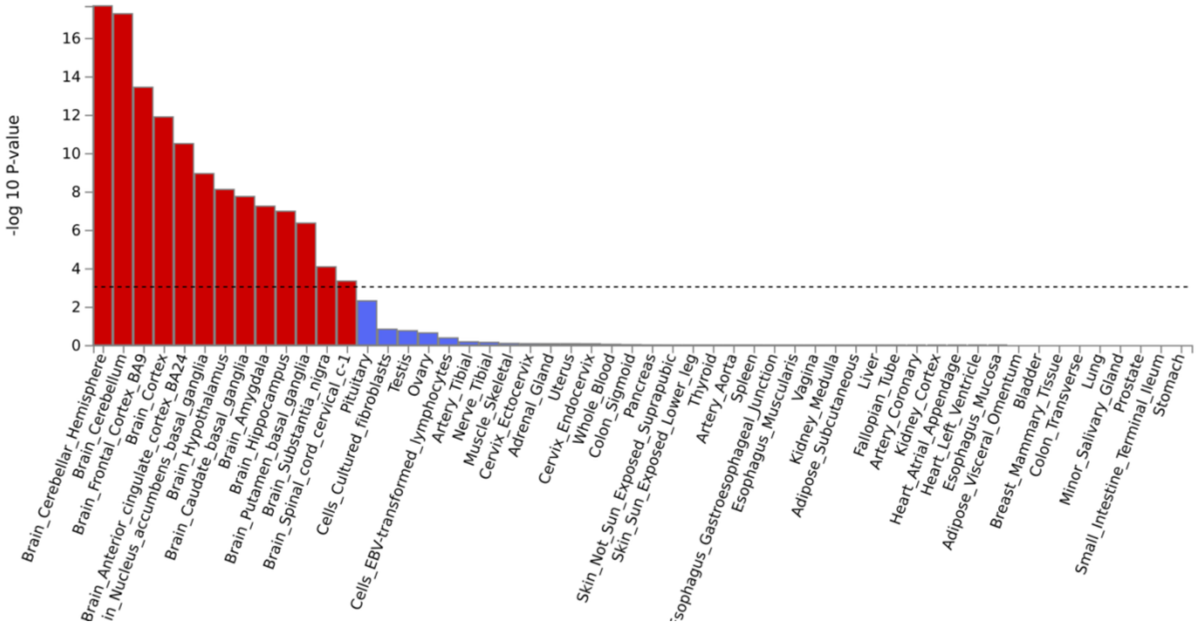


**Supplementary Figure 2.** MAGMA tissue expression with GTEx v8 tissue types, ordered by p-value for EUR internalizing.
